# Enzymatic profiling of cfDNA methylation for detection and monitoring of lung cancer

**DOI:** 10.1101/2025.05.08.25327181

**Authors:** Abed Agbarya, Noa Gilat, Yael Michaeli, Jasline Deek, Assaf Grunwald, Suheil Artul, Rasha Khoury, Michael Peled, Yuval Ebenstein

## Abstract

We present a highly sensitive, low-cost approach for detecting lung cancer and monitoring response to therapy, based on sequencing-free detection of methylation biomarkers in cell-free DNA. An engineered methyltransferase is used to fluorescently label CpG sites. When applied to bisulfite-treated, PCR-amplified cell-free DNA, fluorescent reporters attach to all originally methylated sites, which can then be read on a standard hybridization microarray. In a proof-of-concept study involving 60 blinded participants, we distinguished cancer patients from healthy individuals with both sensitivity and specificity exceeding 90 %.

## Main Text

Lung cancer remains the foremost cause of cancer-related mortality globally^1^. The use of low-dose CT (LDCT) for early detection has shown considerable promise in lowering death rates, as evidenced by studies like the National Lung Screening Trial (NLST)^2^ and the Dutch-Belgian Lung Cancer Screening Trial (NELSON)^3^. However, a major issue with LDCT screening is that numerous, mostly benign, nodules are seen in the lungs during screening, leading to high incidence of false-positive results, which can lead to unnecessary biopsies or surgeries. For example, the NLST reported a 26.3% false-positive rate for initial screenings^4^, while the NELSON trial noted a 19.8% rate. In addition, detected lung nodules smaller than 2 cm pose significant challenges for effective biopsy^5^ and are usually monitored without intervention. Consequently, tumours may develop resistance or metastasize during this period^6^.

Beyond LDCT screening, liquid biopsies may detect biomolecular signatures, offering potential insights into disease status. Tumour-informed tests such as the recently approved Signatera™ lung cancer test^7^, already achieve remarkable sensitivity for relapse and residual disease detection. However, they require a biopsy to determine tumour-specific mutations. When the tumour is not available, cell-free DNA methylation is emerging as a promising biomarker, exhibiting unique signatures at multiple genomic regions in lung cancer patients compared to healthy individuals, and demonstrating significant diagnostic value^8^. Nonetheless, many current liquid biopsies lack the sensitivity required for reliable identification of early-stage or low tumour burden cancers. A more tangible entry point for liquid biopsy is in combination with imaging as a molecular complement. The combined evaluation of CT and cfDNA methylation features in lung cancer cases has not been extensively studied. However, recent successful applications of such multimodal data have been used to assess the staging and risk prediction capabilities of cfDNA methylation sequencing combined with imaging^9, 10^. The need for a facile and complementary molecular test is emphasized considering the low adherence to LDCT screening which is currently on the order of ∼60%^11,12^.

Current and emerging DNA methylation-based tests rely on DNA sequencing. However, the sample enrichment and preparation procedures, the depth of sequencing needed for accurate classification, and the cumbersome computational analysis render these tests inherently expensive. Given these facts, the cost-benefit of such tests for screening or assisting LDCT is unclear^13^. Here we present a newly developed methylation microarray that shows potential for highly sensitive yet low-cost detection of lung cancer from plasma-derived cell-free DNA.

This study enrolled 103 participants (Table S1) with available preoperative blood plasma samples for cfDNA extraction and analysis. Among them, 51 patients were diagnosed with stage 2-4 lung cancer and 52 were age matched controls. A simple hybridization microarray was used to classify the cohorts by their unique methylation signatures.

This is enabled by a novel DNA methylation detection chemistry, a chemoenzymatic reaction that introduces a fluorescent reporter to every unmodified methylation site (CpG) in DNA^14–16^. We hypothesized that this may serve to label all positions of methylated CpG sites in bisulfite treated, PCR amplified cell-free DNA. Since only methylated CpG sites in the original sample become unmethylated CpGs in the amplified sample, only the originally methylated sites are fluorescently labeled by the enzyme. When applying such labeled DNA to a conventional microarray, the fluorescence intensity from each scanned feature reports on the degree of methylation in the particular genomic target defined by the sequence of the capture probes on the array. As proof of concept, we have used a commercially available comparative genomic hybridization array (CGH) from Agilent technologies, designed for copy number analysis. As such, it sparsely covers the entire genome at ∼40kb intervals and is denser in gene bodies and regulatory elements with a total of about 60k genetic loci represented on the array surface. Fluorescently labeled cfDNA samples were prepared for hybridization (see methods), applied to the arrays and scanned after overnight hybridization. Following data normalization, an analytical pipeline was built to identify distinctive methylation loci. For training, we used data acquired from 22 lung cancer samples and 21 healthy controls. When analyzing the methylation patterns, we identified a set of 170 genomic regions with highly differential methylation levels that allowed us to distinguish the two populations with high accuracy. The experimental and analytical workflow are illustrated in figure 1 a-f and in supporting figure 1.

**Figure 1.**
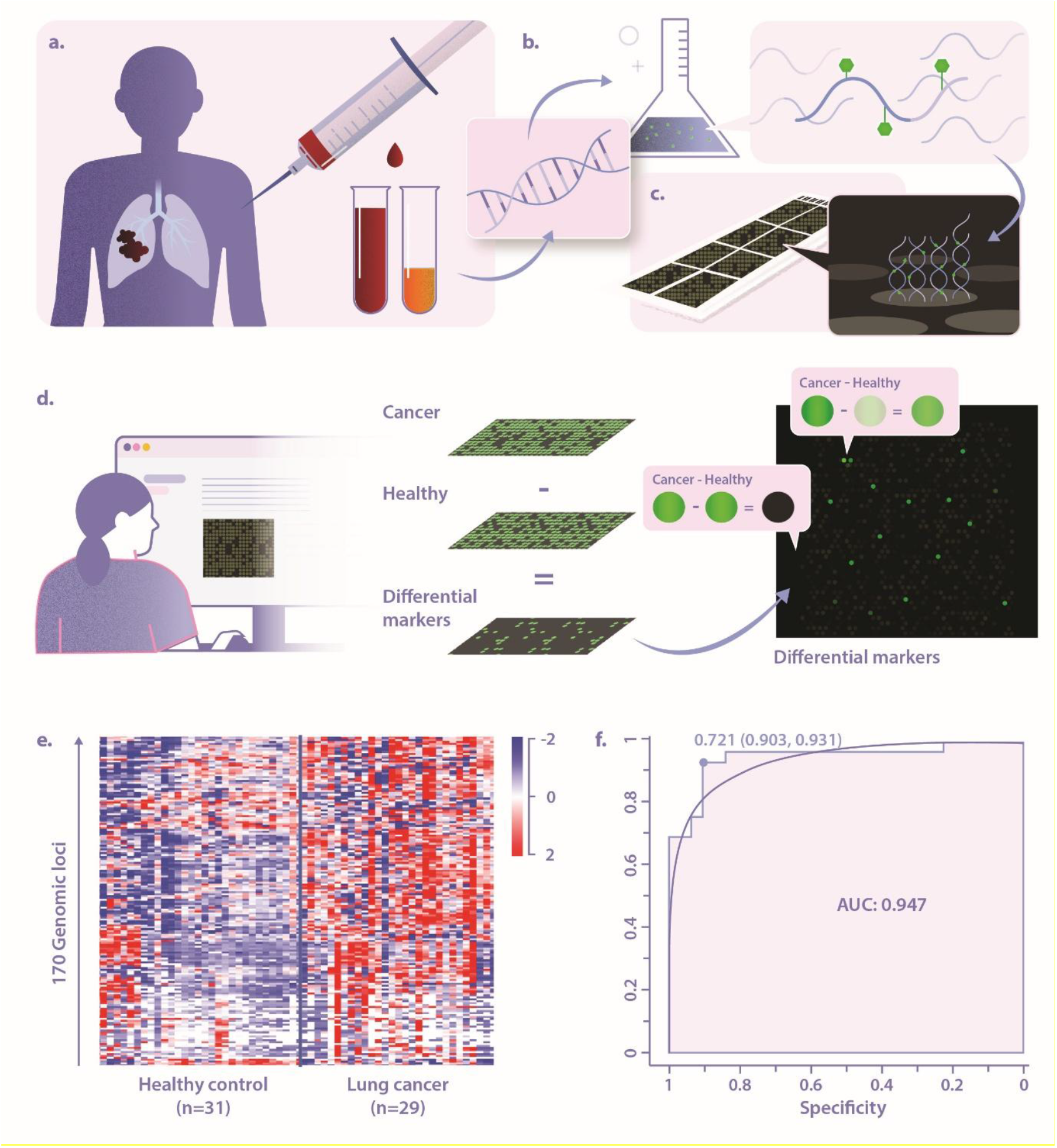
Schematic workflow and classification results. **A**. whole blood is collected and plasma is separated. **B**. Cell-free DNA is extracted from plasma, processed, and fluorescently labeled at methylation sites. **C**. Labeled DNA is applied to a hybridization microarray, directly reporting on the methylation status of every genomic region represented on the microarray. **D**. Comparing the signature of the lung-cancer patients against the controls leaves a small subset of differential regions. **E**. A heatmap showing the methylation levels of the top 170 differential regions in lung cancer and healthy controls. **F**. Receiver operating characteristic (ROC) curve reflecting the classification performance on 60 blinded participants.

Next, a new set of 29 lung cancer samples and 31 controls were blindly classified using the selected biomarker panel. The heatmap in Figure 1g displays the top 170 differential regions across 60 samples tested, showing high contrast between the two populations. ROC analysis showed peak performance of 93.1% sensitivity at 90.3% specificity, with an AUC of 0.947 (Figure 1h).

Our Non-Small Cell Lung Cancer (NSCLC) cohort was composed of two histologic subtypes, adenocarcinoma (N=32) and squamous cell carcinoma (N=17). Independent comparison of the squamous cell signatures to 22 age matched adenocarcinoma patient signatures, revealed a set of 168 methylation markers (not included in the cancer/healthy classifier) that distinguishes between these histological subtypes with an AUC of 88.1%, correctly classifying 86.4% of squamous cell carcinoma samples, and 88.2% of adenocarcinoma samples (Figure S2).

Next, we looked for mechanistic validations that our selected differential panel is biologically associated with lung cancer. Since many of our biomarkers reside outside gene coding regions, we used GREAT^17^ to generate a list of associated genes through proximity or functional interpretation of *cis*-regulatory elements. Among the genes identified were several established oncogenic drivers and tumor suppressors in lung cancer such as BRAF^18^, CDKN2B^19^, SMARK^20^ and RICTOR^21^, as well as several EMT/metastasis regulators such as SNAI1^22^, Vimentin^23^, ROCK^24^,TRIB2^25^ and FYN^26^. We then used DAVID^27^ for signaling pathway enrichment. Stringent analysis yielded two highly significant pathways, Developmental protein (UniPort: KW-9996 & KW-0217) and cyclic adenosine monophosphate (cAMP) signaling (UniPort: hsa04024). Both pathways are directly involved in many cancers and specifically in lung cancer. The developmental module, that contains WNT / β-catenin signaling, contains dozens of genes from our biomarker list, including TCF7L2 (alias TCF-4) which known to modulate WNT signaling in lung cancer^28^. Other markers were related to the cAMP signaling pathway in lung cancer, most notably GNAS, whose hotspot R201C/H mutations characterize a distinct subset of mucinous and non-mucinous lung adenocarcinomas, and VIPR1 (alias VPAC1) which is over-expressed in 58 % of NSCLC and SCLC tumours^29^ (see supplementary information and supplemental file 1).

Finally, we tested whether our assay reports response to therapy as a molecular complement to radiology. We followed two cases of advanced stage NSCLC requiring an aggressive chemo-immunotherapy (9LA) protocol. The patients were tested for cfDNA methylation before and after treatment, alongside chest imaging. Good correlation was observed between the clinical response evaluated by CT or PET-CT and the methylation levels in the differential loci (Figure 2). Patient 1 did not respond to the treatment and showed slight increase in tumor burden (Figure 2. A-B). When placed on a PCA plot based on the patient methylation array data, slight regression is observed. This patient did not exhibit significant change in his methylation profile (Figure 2.E). Patient 2, on the other hand, responded well to the treatment. The PET-CT scan shows pathological uptakes in multiple malignant masses of NSCLC in the right lung, that are almost diminished after 4 cycles of treatment (Figure 2. C-D, white circle). When placed the PCA plot, a distinct shift in the methylation signature toward the healthy control cluster is observed (Figure 2.E).

**Figure 2.**
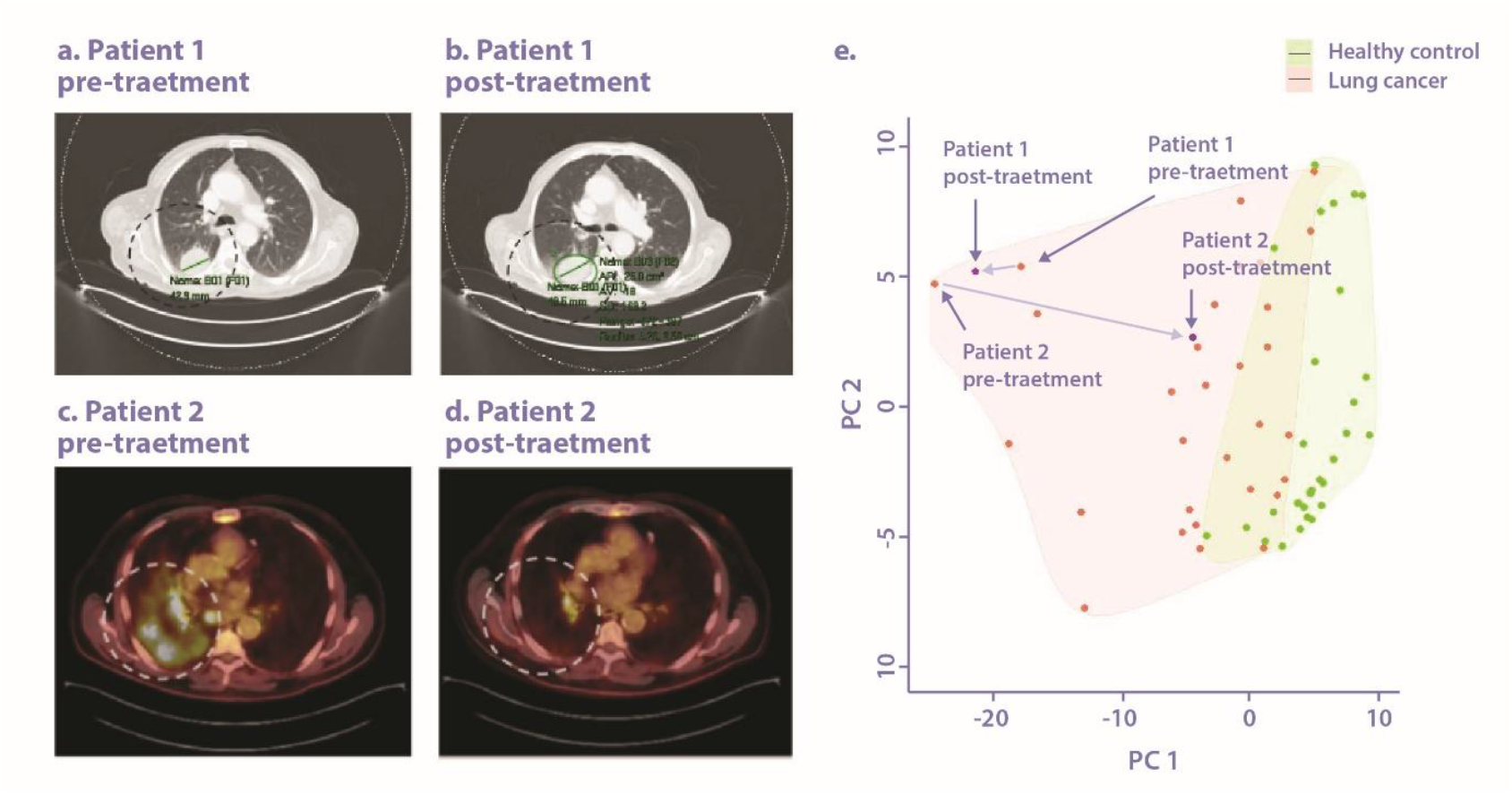
A. The CT scan shows a 42 mm malignant mass (NSCLC) in the Right lung (green section line). B. Tumor Progression to 49 mm after 3 cycles of treatment (Non-Response). C. PET-CT scan shows multiple masses in the right lung (NSCLC) before starting treatment with chemo-immunotherapy. D. Tumor Regression after 4 cycles of treatment (Response). E. PCA plot presenting the distribution of lung cancer (red) and healthy controls (green) samples based on the selected methylation biomarkers. Pink shading represents no false negatives (100% sensitivity), green shading represents no false positives (100% specificity). Pre- and post-treatment samples are overlayed in black. Blue arrows indicate response to therapy.

Thes results presented provide a proof of concept to the potential of this method for complementing chest imaging with molecular information. It may be performed at the point of care in a simple and cost-effective manner, and yielded biologically relevant targets. We note that we have only examined a small fraction of the DNA methylation landscape, limited by the CGH arrays used. Nevertheless, it presents a readily available low-cost alternative to methylation arrays and methylation sequencing. A significant advantage of the proposed approach is its ability to sample a methylation region rather than a specific CpG, resulting in integrated fluorescent intensity from a target region that displays much higher SNR. The methylation atlas has established that CpG methylation occurs in blocks and that these block patterns are cell-type specific^30^. This fact comes to our advantage as it aligns perfectly with the experimental readout of the method. We are currently performing such biomarker discovery on a larger scale, followed by printing a dedicated lung cancer array that will be accessible to the research and clinical community. A larger prospective clinical trial (LUMEN; Eu 101188111) is currently finalizing a protocol for evaluating the test in predicting tumor response to systemic treatment.

*This work was supported by the European Research Council consolidator [grant number 817811]; Israel Science Foundation [grant number 771/21]. Conflict of interest declaration: The IP for this technology has been licensed to a newly formed spinout company, JaxBio LTD. YE is a founder and holds equity in the company, AA is a scientific consultant to the company, YM and NG are currently employed by the company*.

## Methods

The study was conducted in accordance with the Declaration of Helsinki and good clinical practice guidelines following approval by ethics committees and institutional review boards at EMMS (EMMS Hospital Nazareth; Approval number: 37-22-EMMS). All participants provided written informed consent.

### Sample preparation, labeling and hybridization

Whole blood samples were collected in Streck tubes (STRECK). Plasma was separated and cfDNA was extracted with Apostle MiniMax High Efficiency Cell-Free DNA Isolation Kit according to manufacturer’s instructions. Following DNA extraction, adapters were ligated to cfDNA using NEBNext Ultra II End Prep (New England Biolabs) and custom adapters, following with bisulfite conversion using EZ-DNA methylation gold kit (Zymo Research) and PCR amplification, as detailed in the supplementary information.

### Labeling

The “magic sauce” enabling this assay is the use of a bacterial CpG methyltransferase (M.MpeI) that was mutated to allow the transfer of an azide residue to every CpG in the amplified sample^16^. A fluorophore is clicked on to the azide resulting in light emission from every site that was methylated in the original cfDNA (unmethylated CpGs will turn to TpGs and will not be labeled). When applying the sample to a microarray, each array locus emits light in proportion to the amount of methylated CpGs in the original cfDNA sample, generating a unique light intensity pattern that represents the underlying sample methylome. The assay is inherently targeted via hybrid capture on the microarray surface, resulting in an extremely simple and low-cost test to perform and analyze.

Bisulfite-converted and amplified cfDNA samples underwent fluorescent labeling process using mutated CpG methyltransferase. The labeling process resulted in a fluorophore attached to every CpG site that was methylated in the original cfDNA.

The labeled cfDNA samples were hybridized to 60K Human CGH Microarray (SurePrint G3, Agilent Inc.), overnight at 48°C. Following hybridization, the microarray was washed and scanned using a standard slide scanner (Innoscan 1100, Innopsys Inc.). The intensity of each feature on the microarray was measured using the dedicated scanner software (Mapix, Innopsys Inc.).

## Supporting information

Gene list for pathway analysis

## Data Availability

All data produced in the present study are available upon reasonable request to the authors

## Data analysis

A custom analytical pipeline was developed to enable the simultaneous processing of a large number of datasets. The software receives data from healthy and lung cancer samples which serve as a training set. Post normalization, the software identifies the differential markers between healthy and lung cancer samples. The pipeline integrates statistical analysis and feature selection to identify a subset of probes with high discriminatory power for classifying samples as healthy or lung cancer. A new set of samples was introduced to the trained module and blindly tested to classify each sample as either healthy or lung cancer. Specificity and Sensitivity were calculated according to the tested data. A similar process was applied to identify markers that differentiate between lung adenocarcinoma and squamous cell carcinoma.

