## Supporting information for "Enzymatic profiling of cfDNA methylation for detection and monitoring of lung cancer"

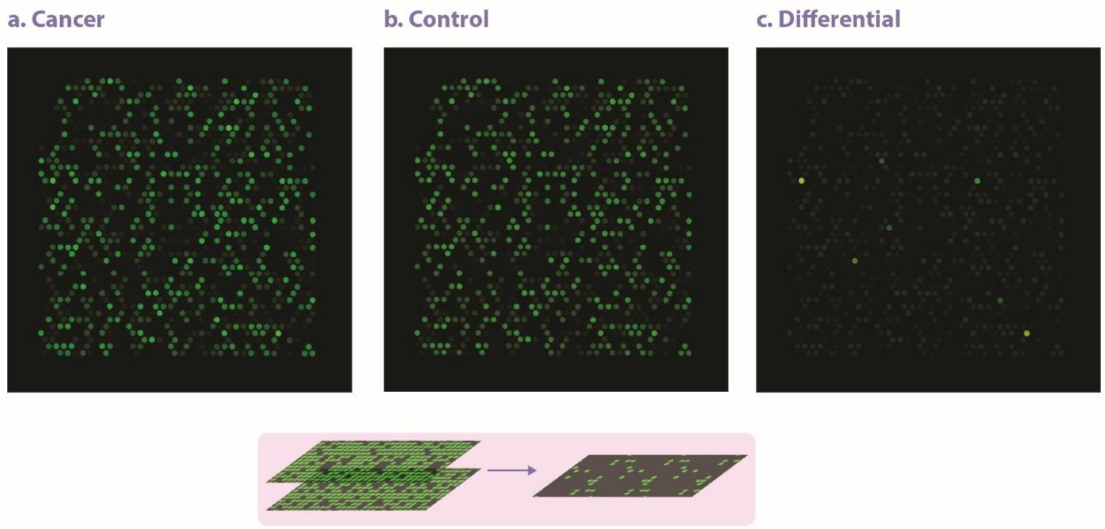


**Figure S1-** Raw normalized image scans of the same ~ 1200 feature region in two microarrays: (a). Lung cancer and (b.) healthy control. Each feature on the array targets cfDNA from a specific genomic locus. Signal intensity is proportional to the degree of methylation in this locus. A differential image (c.) shows only features that display significant detectable differences between the methylation levels in the lung cancer VS. the healthy control. This small subset of targets are later used to classify lung cancer samples VS. controls.

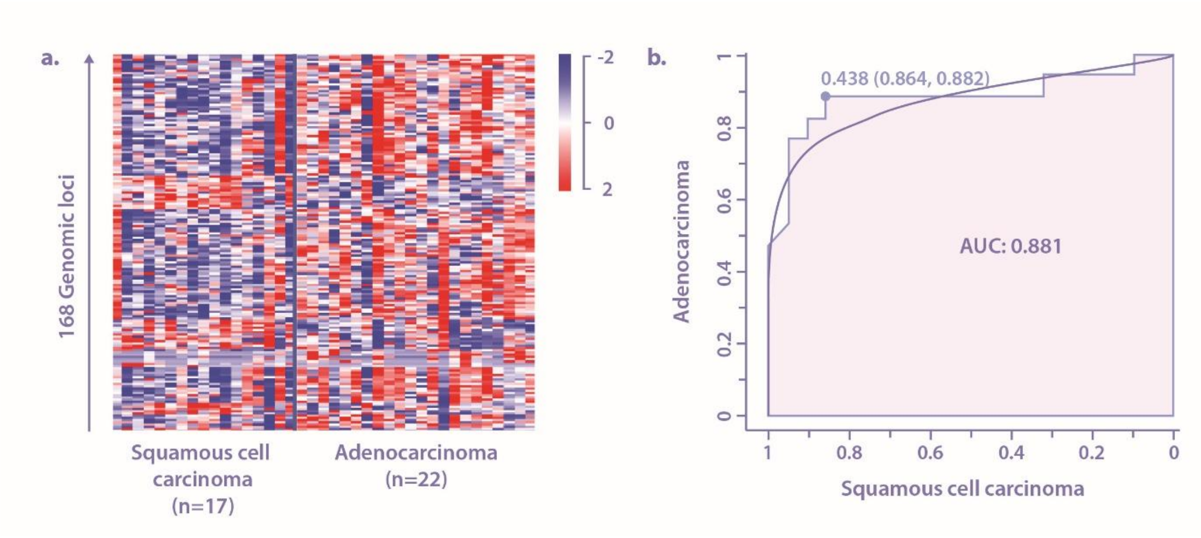


**Figure S2-** Classification of different subtypes of lung cancer. A. Heat map displaying 168 differential methylation biomarkers that classify two subtypes of lung cancer, squamous cell carcinoma and adenocarcinoma. B. ROC curves presenting the classification performance of squamous cell carcinoma and adenocarcinoma.

**Table S1**

| | | Lung Cancer | | | Healthy |
|---|---|---|---|---|---|
| | Lung Cancer Subtype | Adenocarcinoma | Squamous cell carcinoma | Unknown subtype | |
| Age | Age Median(IQR) | 61.5(12.75) | 62(8) | 61.5(8.5) | 61(10.5) |
| Gender | Male | 24 | 15 | 1 | 30 |
| | Female | 8 | 2 | 1 | 22 |
| Smoking | Yes | 22 | 10 | 0 | 1 |
| | no | 10 | 7 | 0 | 16 |
| | NA | 0 | 0 | 2 | 35 |
| Disease stage | IIB | 2 | 3 | 0 | N/A |
| | IIIA | 5 | 1 | 0 | N/A |
| | IIIB | 1 | 2 | 1 | N/A |
| | IIIC | 2 | 1 | 0 | N/A |
| | IV | 3 | 0 | 1 | N/A |
| | IVA | 14 | 7 | 0 | N/A |
| | IVB | 5 | 3 | 0 | N/A |
| Total | | **32** | **17** | **2** | **52** |

**Table S1: Clinical and demographic characteristics of study participants. Summary of age, gender, smoking status, and disease stage for lung cancer patients (stratified by subtype) and healthy controls.**

**Supplementary methods**

**Sample Preparation**

Whole blood samples (4-8 mL) were collected in Streck tubes (STRECK) to ensure cfDNA stabilization. Plasma was separated by centrifugation in two steps: first at 1600g for 10 minutes, followed by careful transfer of the upper plasma layer to a new 15 mL tube without disturbing the buffy coat. A second centrifugation was performed at 5000g for 10 minutes to further purify the plasma. cfDNA was extracted using Apostle MiniMax High Efficiency Cell-Free DNA Isolation Kit according to the manufacturer's instructions. Following extraction, cfDNA underwent adapter ligation using the NEBNext Ultra II End Prep kit (New England Biolabs) with custom adapters to prepare for downstream processing. Bisulfite conversion was performed with the EZ-DNA methylation Gold kit (Zymo Research) to enable differentiation between methylated and unmethylated cytosines. The converted cfDNA was amplified via PCR under the following conditions: 95°C for 2 minutes, followed by 18 cycles of 95°C for 15 seconds, 55°C for 15 seconds, and 72°C for 30 seconds. A final extension was performed at 72°C for 2 minutes, and samples were held at 12°C indefinitely. PCR products were cleaned using AMPure beads (bead ratio 1.2x, 60 µL), and DNA was eluted in 30µl for downstream labeling and hybridization.

**Fluorescent Labeling of cfDNA**

The key innovation of this essay is in the chemoenzymatic labeling of methylated CpG sites. A mutated bacterial CpG methyltransferase uses a synthetic cofactor to transfer an azide residue to every CpG site in the amplified cfDNA sample. This enzyme selectively targets methylated CpGs, since unmethylated CpGs are converted to TpGs during bisulfite treatment and amplification and are thus not recognized. Following the enzymatic transfer, a fluorophore was covalently attached to the azide residues using click chemistry. The labeling reaction was performed according to Avraham et al[1].

**DNA Microarray Hybridization**

Following fluorescent labeling, 500 ng of the labeled cfDNA amplicons were hybridized onto Agilent 60K Human CGH microarrays (Agilent Technologies). Pre-hybridization solution was prepared in a 50 mL tube by combining 0.5 g BSA (Sigma), 41 mL double-distilled water (DDW), 8.75 mL SSC 20x, and 250 µL SDS 20%. Agilent microarray slides were incubated in the pre-hybridization solution at 37°C for 20 minutes. Labeled cfDNA samples were prepared with 2X Agilent hybridization buffer. A total volume of 100 µL was prepared by mixing 50 µL of labeled DNA with 50 µL of Agilent 2X hybridization buffer. Samples were incubated at 98°C for 3 minutes and were then incubated at 37°C for 30 minutes before immediate hybridization onto the Agilent array. Hybridization was performed overnight at 48°C with mild rotation to ensure efficient and uniform binding of cfDNA to complementary probes on the array surface. Post-hybridization, the microarrays were washed twice with PBST for 1 minute to remove non-specifically bound DNA. The slides were scanned using the Innoscan 1100 scanner (Innopsys), and fluorescence signal intensities were background subtracted and quantified with the Mapix software (Innopsys).

**Data Analysis**

The analytical data pipeline consists of two main modules: preprocessing and training with feature selection. This pipeline is utilized in both the training and testing.

First, the system processes epigenetic data extracted from the microarrays, systematically eliminating extraneous information during both training and testing stages.

(i) Preprocessor Module:

To address potential biases arising from data heterogeneity, array-specific variations, or differences in sample conditions, robust preprocessing procedures are applied consistently during both the training and testing stages. The initial step in preprocessing involves normalizing the array fluorescence intensity data.

The normalization strategy leverages the observation that over 99% of methylation sites on the CGH arrays are consistent across all arrays and samples, and it relies on the internal calibration probes embedded within the arrays. After evaluating several approaches, polynomial fitting was identified as the most effective normalization method, successfully producing uniform illumination levels across all slides. This process achieved over 99% similarity between any two compared samples. To minimize batch effects, each slide was loaded with an equal number of cancer and healthy samples. Furthermore, multiple experiments were conducted across different slides and at different times to differentiate true biological signals from technical variability. To ensure robustness, both the training set and the test set were composed of a diverse selection of samples originating from all slides.

(ii) Training and Feature Selection Module:

To identify differential methylation markers distinguishing healthy from lung cancer samples, a statistical T-test was performed with a significance threshold of $p < 0.01$. In addition, only features showing a differential methylation ratio greater than 20% between the two groups were selected, resulting in a final set of 150–200 features capable of discriminating between healthy and lung cancer samples. A training cohort comprising 21 healthy and 22 lung cancer cfDNA samples was used for marker discovery and model training. A simple classification process was followed. The methylation level of every biomarker in the tested sample is compared to the average methylation level of the same feature in the training cohorts. A value of 1 is given to this feature if it is closer to the healthy methylation level and a weight of 0 if it is closer to the lung cancer methylation value. Next, for each sample, a score is calculated by summing all the assigned values (of all selected methylation features), with each feature contributing equally; samples are classified as lung cancer or healthy based on whether their score falls above or below a predefined threshold.

Subsequently, an independent validation set of 29 lung cancer and 31 healthy cfDNA samples, not included in the training phase, was analyzed to evaluate the model’s performance. Based on their methylation profiles, each sample was assigned a score classifying it as either healthy or lung cancer. Specificity and sensitivity were calculated from the blind-testing results, providing a quantitative assessment of the assay’s diagnostic accuracy.

**Pathway analysis**

Since most of our 155 differential sites did not reside within gene bodies, we utilized the GREAT web tool (ref 17) to identify genes associated with these markers. GREAT analysis was performed using the "Basal plus extension" threshold mode, configured to include regions constitutively 5.0 kb upstream and 1.0 kb downstream of genes, with an additional extension of up to 1000.0 kb permitted when markers were located between genes and associated enhancers. This analysis yielded 265 genes linked to our differentially methylated markers.

To identify enriched cellular pathways, the list of associated genes obtained from GREAT was analyzed using the DAVID web tool (ref 27). Resulting pathways were filtered to retain only those with an adjusted p-value (Benjamini correction) below 0.05. All reported pathways exhibited a fold enrichment greater than 2. The table below shows groups of genes associated with our biomarker panel with peer reviewed evidence of involvement in lung cancer (driver mutations, focal amplifications/deletions, prognostic biomarkers, functional data in cell/animal models).[2]

| **Category** | **Gene(s)** | **Nature of the evidence** |
|---|---|---|
| **Established oncogenic drivers / tumour suppressors** | **BRAF**, **CDKN2B**, **SMARCA2**, **RICTOR** | **BRAF** V600E mutations define a targetable NSCLC subset[3]. **CDKN2B** deletions/methylation are frequent in NSCLC and correlate with poor outcome[4]. **SMARCA2** loss is lethal in SMARCA4-mutant lung cancers, opening a degrader/PROTAC strategy[5]. **RICTOR** amplification (~13 % of LUAD) activates mTORC2 signalling; considered actionable[6]. |
| **EMT / invasion / metastasis regulators** | **SNAI1**, **VIM (Vimentin)**, **ROCK1**, **ROCK2**, **TRIB2**, **FYN** | Snail (**SNAI1**) up-regulation worsens prognosis in lung SCC[7]. **Vimentin** overexpression may predict the progression and an unfavorable survival of NSCLC[8]. **ROCK1/2** signaling and **TRIB2** or **FYN** over-expression promote migration and metastasis[9–11]. |
| **Recurrent (but lower-frequency) mutations / expression changes** | **RUNX1**, **Ki-67**, **BMPR1B**, **KIF18A**, **NPNT** | **RUNX1** alterations associate with shorter survival in LUAD[2]. High **Ki-67** labelling is an adverse prognostic factor in early-stage NSCLC. Blocking **BMPR1B** (ALK6) triggers apoptosis and suppresses clonogenic growth of multiple NSCLC lines[12]. **KIF18A** over-expression correlates with higher stage and poorer overall survival in NSCLC[13]. |
